# Disrupted Coupling of Heart Rate–Dependent Brain Network Switching and Attentional Task Performance in Schizophrenia Spectrum Disorders

**DOI:** 10.64898/2026.04.06.26350241

**Authors:** Kimberly Kundert-Obando, Andrew R. Kittleson, Shiyu Wang, Haatef Pourmotabbed, Ella Provancher, Anna Machado, Baxter P. Rogers, Sohee Park, Julia M. Sheffield, Heather B. Ward, Catie Chang

**Author notes:** **Correspondence: Kimberly Kundert-Obando,**, **Catie Chang,**, **Heather Ward,**, **Julia Sheffield,**, 400 24th Avenue S, Nashville, TN 37240.

## Abstract

Cognitive deficits are a core feature of schizophrenia, yet their neural mechanisms remain poorly understood. Network switching—a measure of how frequently brain networks change their interactions over time—has been linked to cognitive performance in healthy individuals and has been reported to be altered in schizophrenia. Recent evidence further suggests that the relationship between network switching and cognition depends on arousal, which is itself disrupted in schizophrenia. However, whether arousal-related alterations in network switching contribute to cognitive impairment in schizophrenia remains unclear. Here, we used concurrent resting-state functional MRI (fMRI) and pulse oximetry data from 39 healthy controls (HC), 27 psychiatric controls (PC), and 39 individuals with schizophrenia spectrum disorders (SSD) to examine whether network switching relates to indices of autonomic arousal. Additionally, in HC and SSD participants, we tested whether arousal moderated the association between network switching and performance on an attention task. We observed no group differences in autonomic arousal. However, PC exhibited higher dorsal default mode and anterior salience network switching rates compared to SSD participants. Additionally, autonomic arousal significantly moderated the relationship between network switching and cognitive performance in HC, an effect that was absent in SSD. Notably, these findings implicate network switching as a potential neural biomarker that differentiates PC from SSD. They also suggest that disrupted coupling between arousal state and network switching, rather than switching alone, may underlie cognitive dysfunction in SSD.

## Introduction

Schizophrenia is characterized by an altered experience of reality, including positive symptoms (e.g., delusions and hallucinations), negative symptoms (e.g., blunted affect and avolition), and cognitive impairment. Despite treatment, it is estimated that 80% of patients with schizophrenia continue to exhibit cognitive impairment that worsens over time (McCleery & Nuechterlein, 2019). Cognitive impairment plays a pivotal role in recovery outcomes, including the ability to obtain and maintain employment and to engage socially (Mascio et al., 2021; McCleery & Nuechterlein, 2019). Therefore, there is a critical need to identify neural markers that can inform the development of targeted interventions for cognitive impairment in schizophrenia.

Switching rate is a neural marker that quantifies the number of transitions of brain regions or large-scale networks between communities over a given period. Research has identified switching to be associated with a range of cognitive functions, including learning, working memory, attention, and executive function (Bassett et al., 2011; Betzel et al., 2017; Braun et al., 2015; Dimitriadis et al., 2021; Pedersen et al., 2018; Shine et al., 2016). Notably, all of these cognitive functions are also impaired in schizophrenia (McCutcheon et al., 2020). Although abnormal switching rates of brain regions and large-scale networks have been identified in schizophrenia (Braun et al., 2015; Gao et al., 2024; Gifford et al., 2020; Hu et al., 2025; Wei et al., 2022; Zhen et al., 2025), only two studies have examined their relationship with cognition (Braun et al., 2015; Hu et al., 2025). Braun et al. reported a higher global (i.e., average) switching rate in participants with schizophrenia during a working memory task, and Hu et al. 2025 found that, at rest, switching rates of the anterior cingulate cortex and parahippocampal gyrus were related to impairments in problem-solving, attention, and working memory. The anterior cingulate cortex is a key region of the salience network, while the parahippocampal gyrus is part of the default mode network (Buckner et al., 2008; Seeley, 2019). Thus, although switching dynamics in both the salience network and the default mode network are known to be abnormal in schizophrenia, it remains unclear whether these alterations are related to cognitive impairments, including, for example, attention.

We previously established that, in healthy controls, switching rates for both the anterior salience network and the default mode network depended on arousal state (Kundert-Obando et al., 2026). Specifically, increased switching of the anterior salience network and reduced switching of the default mode network occurred during high arousal (alert wakefulness) compared to low arousal (drowsiness and light sleep). In individuals with schizophrenia, both elevated salience network switching (Braun et al., 2015) and reduced default mode network switching (Gifford et al., 2020; Li et al., 2025) have also been reported, suggesting that heightened arousal might drive abnormal dynamics in brain switching rates in schizophrenia. In support of this hypothesis, arousal is known to be heightened in individuals with schizophrenia, as reflected in autonomic arousal measures such as higher mean heart rate and lower heart rate variability (Benjamin et al., 2021; Clamor et al., 2019; Datta et al., 2023; Schlier et al., 2019). Additionally, heart rate variability is found to be lower in multiple psychiatric disorders (Gorman & Sloan, 2000; Ramesh et al., 2023), and switching rate patterns have been found to differentiate bipolar disorder and major depressive disorder (Han et al., 2020). However, it is unknown whether altered autonomic arousal in these clinical populations might contribute to altered switching rates. Further, although we have found that arousal state moderates the relationship between global brain network switching and cognitive performance in healthy controls, it is unknown if this phenomenon extends to individuals with schizophrenia. We hypothesize that these arousal-network interactions will be disrupted in participants with schizophrenia-spectrum disorders, unveiling a potential target for reducing cognitive impairment in schizophrenia.

To test this hypothesis, here we leveraged concurrent resting-state fMRI and pulse oximetry data to investigate interactions among brain network switching, autonomic arousal, in a group of healthy controls (HC), psychiatric controls (PC), and participants with schizophrenia spectrum disorder (SSD). We focused on the anterior salience network along with the dorsal and ventral subdivisions of the default mode network, as their dynamics were previously shown to be modulated by arousal (Kundert-Obando et al., 2026). We first tested whether the switching rates of these networks are altered across these three groups. Since switching rate was found to differentiate those with bipolar disorder and major depressive disorder (Han et al., 2020), we hypothesized our PC participants would have different switching rates in comparison to SSD participants. Next, we examined how these dynamics related to autonomic arousal and whether this relationship differed across our three groups. To probe broader alterations in brain dynamics, we also assessed the global switching rate and its association with attention as one component of cognition. Finally, we tested whether autonomic arousal modulated the relationship between cognitive performance in an attentional task and switching rate (both global and network-specific), and whether such modulatory effects were disrupted in schizophrenia spectrum disorder.

## Methods

### Participants

This study drew upon two datasets for a combined total of 75 participants with schizophrenia spectrum disorder (SSD) and 76 controls (46 healthy controls [HC]and 30 psychiatric controls [PC]). Both datasets were acquired at Vanderbilt University Medical Center using similar recruitment procedures, and psychiatric diagnoses were established through a Structured Clinical Interview for DSM-5 Disorders (SCID) (First, 2015). SSD participants in this study were diagnosed with schizophreniform disorder, schizophrenia, or schizoaffective disorder, and did not have any comorbid neurological disorders or history of significant head trauma. HC participants had no history of diagnosed psychiatric disorders. Individuals in the PC group had no lifetime history of psychosis, confirmed by DSM-5 SCID interview, but could meet criteria for other psychiatric disorders, including depression, anxiety, attention-deficit hyperactivity disorder, and bipolar II disorder. Overall, controls (PC and HC) for both studies were excluded if they had a history of psychosis. All participants provided written informed consent in accordance with the institutional review boards at Vanderbilt University Medical Center.

### Data Acquisition

A total of 41 SSD, 39 HC, and 29 PC had completed an MRI scan with simultaneous physiological recording via photoplethysmography (PPG). In the first dataset, a total of 21 SSD and 39 HC completed a high-resolution anatomic scan (TFE sequence, TR/TE = 8.0/3.7 ms, FA = 8°, voxel size = 0.7mm isotropic, FOV = 256 mm x 256 mm) and one resting-state fMRI scan (EPI sequence, TE= 35 ms, TR = 1300 ms, duration of scan= 8 min, voxel size = 2.5 mm isotropic, FA = 79°, FOV = 240 mm x 240 mm) with simultaneous PPG and were instructed to keep their eyes open while attending to a fixation cross. In the second dataset, a total of 22 SSD and 29 PC completed a high-resolution anatomic scan (TFE sequence, TR/TE = 6.7/3.0 ms, FA = 8°, voxel size = 1mm^3, FOV = 240 mm x 240 mm) and two resting-state fMRI scans taken sequentially (EPI sequence, TE= 28 ms, TR = 2000 ms, duration of scan= 10 min, voxel size = 3.0 mm isotropic, FA = 90°, FOV = 240 mm x 240 mm x 114 mm) with simultaneous PPG and were asked to keep their eyes open during the fMRI scans. After visual inspection of the physiological data for movement or signal retrieval artifacts and fMRI data for movement artifacts (for details see MRI preprocessing), data from a total of 39 SSD, 38 HC, and 27 PC subjects were included in this investigation. As some SSD and PC participants had two resting state scans in the second dataset, the total number of simultaneous fMRI-PPG scans in the full combined sample was 51 SSD, 38 HC, and 46 PC.

### Continuous Performance Task

Of the participants who had adequate MRI and physiological data quality, a total of 39 HC and 21 SSD completed the MATRICS Continuous Performance Task-Identical Pairs (CPT) task outside of the scanner (Rapisarda et al., 2014). This neuropsychological task of sustained attention and vigilance required participants to attend to quickly flashing numbers on a computer screen and respond via button press when consecutive identical numbers were presented. Participants completed three conditions of this task, with two-, three-, and four-digit numbers shown to capture subtle deficits in progressively more difficult levels of sustained attention. Performance was quantified using d’, a sensitivity index that measures the ability to discriminate target stimuli from non-target stimuli, with higher scores representing better discrimination abilities.

### MRI Preprocessing

Anatomical images were segmented into gray matter, white matter, and cerebrospinal fluid compartments with the Computational Anatomy Toolbox 12 (CAT12, version 12.5; http://www.neuro.uni-jena.de/cat/). Resting-state scans were preprocessed in SPM12 (https://github.com/vuIIS/vuiis-cci-info?tab=readme-ov-file#citing-xnatdax), where they were first realigned to a mean volume to reduce head-motion effects, and then coregistered with the native-space structural scan and MNI 152 template. Next, they underwent simultaneous bandpass filtering (0.01–0.1 Hz) and nuisance regression of six head motion parameters and six principal components of CSF and white matter (Hallquist et al., 2013). Code for this preprocessing pipeline, which was applied to both datasets, is available at https://github.com/baxpr/connprep. All resting-state scans went through a quality assurance procedure that included calculating framewise displacement (FD) as a measure of head motion during scanning. Scans with a mean FD>0.5 mm or that had one or more motion artifacts greater than 5 mm were excluded (n=27).

### Deriving Heart Rate and Heart Rate Variability

Heart rate (HR) was derived from the PPG data, band-pass filtered with a second-order Butterworth filter (0.5–2 Hz). Inter-beat intervals (IBIs) were computed from peaks detected with the findpeaks function of MATLAB (minimum height: 5% interquartile range), and transient artifacts were corrected via interpolation of outlier points (more than three standard deviations away from the mean). HR was calculated as the inverse of the mean IBI averaged over the total scan duration (Chang & Glover, 2009; Chen et al., 2020). Additionally, we computed the logarithmic transformation of the mean HR to normalize the variability, which was applied towards the moderation statistical analysis.

Heart rate variability (HRV) was computed using two different methods. First, we computed the root mean square of successive differences (RMSSD) by squaring the difference of two consecutive IBIs, averaging over the entire scan duration, and taking the square root of that value. The second HRV measure was the standard deviation of IBIs (SDNN) over the entire scan duration. To normalize the data, we conducted a logarithmic transformation for both HRV-RMSSD and HRV-SDNN. We included both HRV measures because they provide complementary information about parasympathetic activity in autonomic arousal: RMSSD measures vagal tone, which reflects rapid activity from the parasympathetic system, while SDNN approximates the balance between parasympathetic and sympathetic activity (Gullett et al., 2023; Shaffer & Ginsberg, 2017).

### Deriving Network Switching Rates

Following the methods described in Kundert-Obando et al. (2026), we used an a priori functional atlas (FINDLAB atlas) comprising 14 predefined subnetworks derived from an independent dataset (Shirer et al., 2012). Network time-series were extracted using dual regression (Nickerson et al., 2017). Dual regression is a two-step procedure in which group-level network masks (here, atlas-defined networks) are first spatially regressed onto individual fMRI scans on a volume-by-volume basis to estimate subject-specific network time-series, and these network time-series are then regressed onto the time-series of each voxel to obtain subject-specific spatial maps. Dynamic connectivity was estimated using sliding-window Pearson correlations between all network pairs, with negative correlations set to zero. Window lengths were approximately three minutes with a 1-TR overlap following previous established methods and stability metrics for conducting dynamic analysis in fMRI (Hutchison et al., 2013; Kundert-Obando et al., 2026; Pedersen et al., 2018). Network switching was quantified using an iterative ordinal multilayer Louvain algorithm to track community structure over time (Mucha et al., 2010). Multilayer modularity was computed on a consensus matrix generated from 10 repeated modularity runs, capturing the probability that pairs of networks were assigned to the same community across iterations. Network switching was defined as the proportion of time windows in which a network changed its community assignment (Bassett et al., 2011). For this analysis, we focused on three primary networks--ventral default mode network (VDMN), default mode network (DMN), and anterior salience network (ASAL)--and how they switched across communities that included the 14 subnetworks. Global brain switching was calculated as the average switching across all fourteen subnetworks.

### Statistical Approach

All statistical analyses were completed using RStudio/2024.12.0+467. We tested for group differences in age, sex, race, and nicotine usage in both the full combined sample and the subsample that was used to compare cognitive behavioral measures across groups. A chi-square test was applied to compare gender across groups, and a Fisher’s exact test was used to identify whether distributions of racial and nicotine users significantly differed across groups. Differences in CPZ equivalents between PC and SSD were tested using a one-sample t-test. Group comparisons of age, autonomic arousal (i.e., HR and HRV), and switching rate were carried out using a one-way analysis of variance (ANOVA), with False Discovery Rate (FDR) correction applied across the set of tests performed for a specific measure (e.g., autonomic arousal, switching rate). From visual inspection, our results indicated that mean HC had the greatest difference across groups, and therefore, our following analysis only tested mean HR as our primary autonomic arousal measure.

To examine the effects of mean HR and clinical group on switching rate, we fitted a linear mixed-effects model using the *lme4* package in R. The model included mean HR, group (HC, PC, SSD), and their interaction as fixed effects, with age, gender, CPZ equivalents, and nicotine usage (user or non-user) included as covariates. A random intercept for each participant was included to account for repeated measures within subjects. We replicated the same linear mixed-effects models to test if nicotine usage and clinical group had an effect on switching rate.

For analyses involving the CPT task, only two groups (HC and SSD) were available; therefore, group differences on performance were examined using an independent-samples *t*-test. We focused on an intermediate level of difficulty (three-digit numbers) because task difficulty is inversely related to the threshold for optimal arousal (Anderson, 1994; Broadhurst, 1959). Therefore, easier tasks require higher arousal levels to achieve optimal performance, which may not be reflected in levels of HR captured during fMRI. In contrast, performance at moderate task difficulty may better correspond to the range of arousal levels measured at rest. To test whether switching rate was related to attentional task performance, we conducted a three-way interaction of group, switching rate (global or ASAL), and log-transformed HR on cognitive task performance (CPT∼group*switching*hr_log), controlling for age, gender, and CPZ equivalents. FDR correction was applied across all tested interaction terms in our three-way interaction tests that spanned global switching and anterior salience network models. Secondly, to test whether switching rate relates to attentional task performance for each group individually we constructed two separate linear mixed-effects models (one for HC, one for SSD) while covarying for age, gender, and total CPZ equivalents. FDR corrections were applied within each group analysis (SSD, HC).

## Results

### Group Characteristics Across Group

In the full sample of the data, gender did not significantly differ across SSD, PC, and HC groups (*X*^2^ (2)=1.61, p=0.45). However, there were significant differences in race (p<0.001) and nicotine usage (p<0.001) among groups. A one-way ANOVA revealed a significant effect of group on age, F(2, 101) = 8.87, *p* < .001. Post-hoc Tukey comparisons indicated that the PC group was significantly older than both the HC group (*p* = .001) and the SSD group (*p* < .001). The HC and SSD groups did not differ from one another (*p* = .999). The SSD group had significantly higher antipsychotic doses than the PC group (*t*(47)=4.76, *p* < 0.001).

### Group Differences in Autonomic Arousal

Group differences between HC, SSD, and PC in mean HR and log-transformed HRV measures (RMSSD and SDNN) were assessed. There were no significant group differences (mean HR: F(2,132) = 2.24, q=0.17; HRV log-RMSSD: F(2,132)=1.17, q=0.31; HRV log-SDNN: F(2,132)=2.65, q=0.17; **Figure 1**).

**Figure 1.**
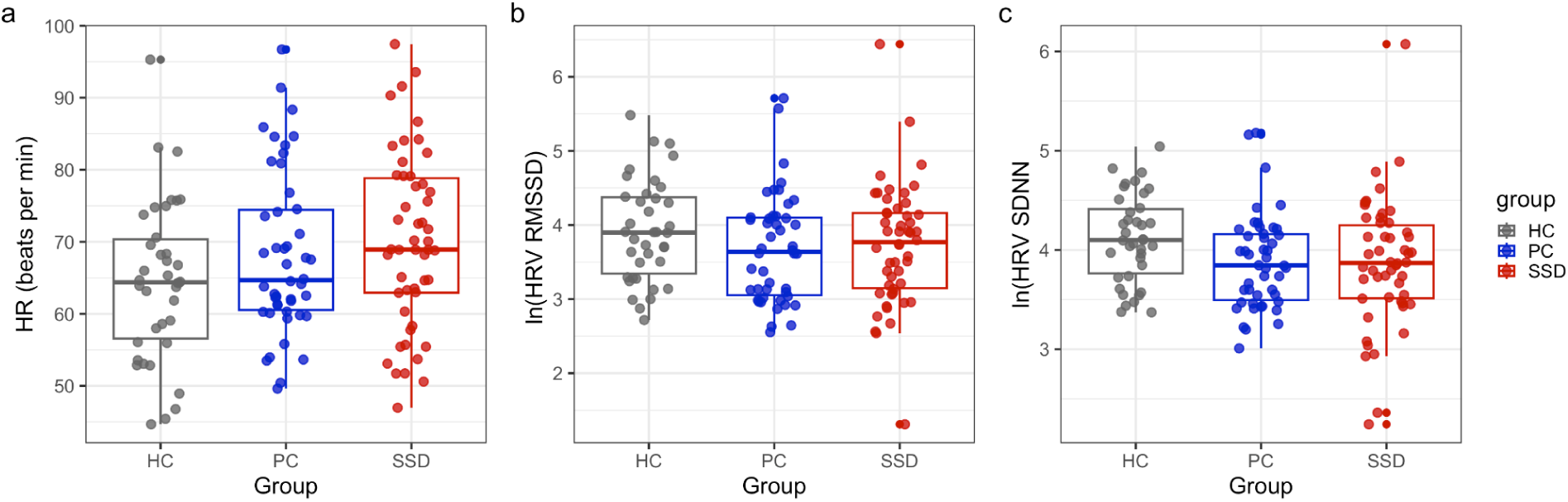
Comparison of autonomic arousal measures between groups. Healthy controls (HC) in grey, participants with schizophrenia spectrum disorder (SSD) in red, and psychiatric controls (PC) in blue. **a)** Mean heart rate (HR), **b)** log-transformed heart rate variability computed through root mean square of successive differences (ln(HRV RMSSD)), **c)** log-transformed heart rate variability computed through standard deviation of inter-beat intervals (ln(HRV SDNN)).

### Group Comparison of Brain Network Switching Rate

We next tested whether the switching rates of the dorsal default mode (DDMN), ventral default mode (VDMN), and anterior salience (ASAL) networks and global switching altered across groups **(Figure 2 a-d)**. We found a significant group difference for ASAL (F(2,132)=6.59, q=0.006), DDMN (F(2,132)=3.89, q=0.03) and Global Switching (F(2,132)=17.05, q<0.001) but not for VDMN (F(2,132)=2.83, q=0.06). Post hoc Tukey comparisons indicated that ASAL switching was significantly higher in PC than HC (p=0.01) and SSD (p=0.004), and DDMN switching was also higher for PC than SSD (p=0.02). Global switching was highest in the PC group compared to both SSD (p=0.003) and HC (p<0.001), and was also higher in SSD compared to HC (p=0.03). Formal statistical results are reported in **Table 2**.

**Figure 2.**
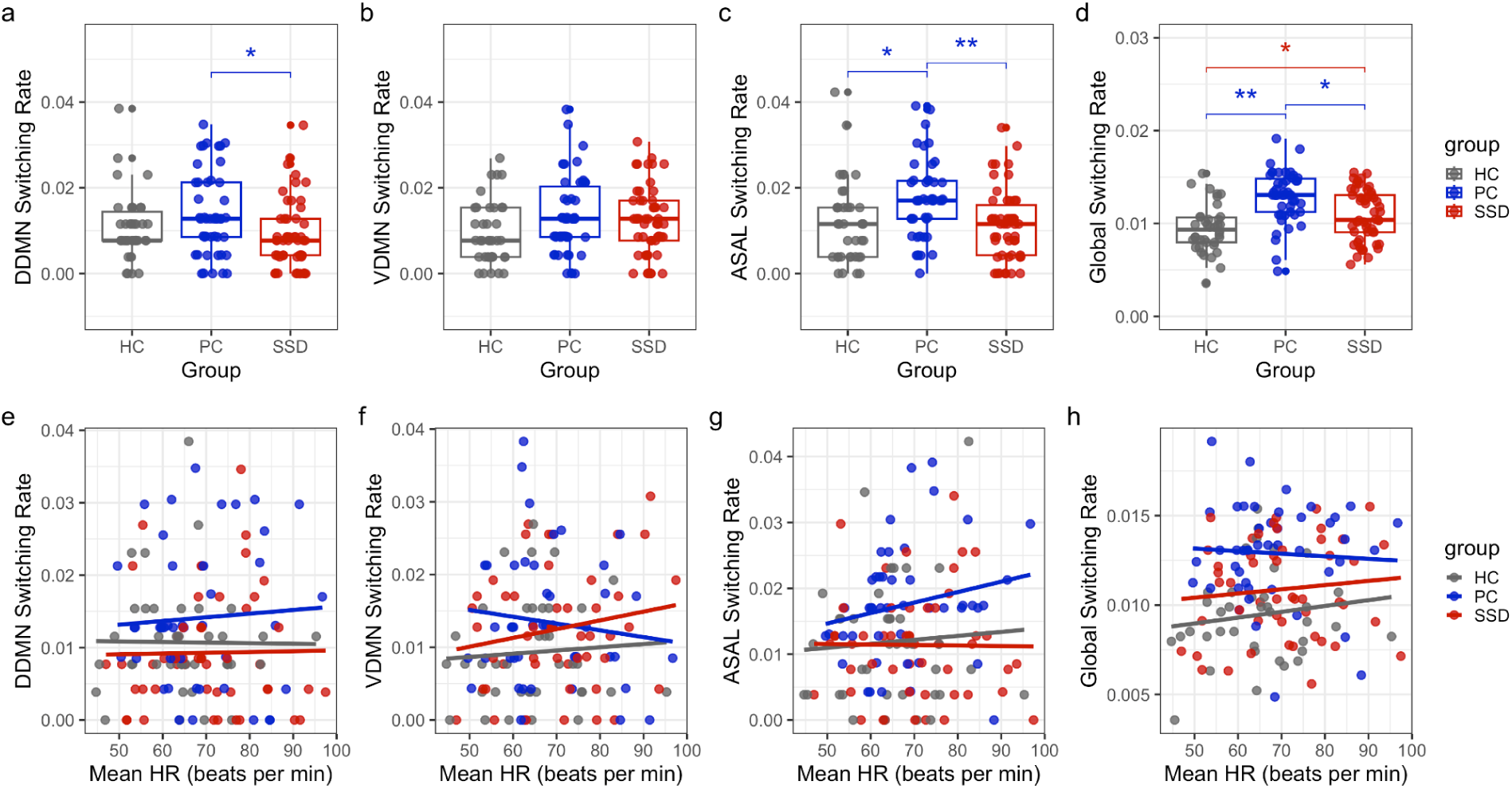
Group Comparison of Switching Rate and Its relation to Mean Heart Rate. Healthy controls (HC) in grey, participants with schizophrenia spectrum disorder (SSD) in red, and psychiatric controls (PC) in blue. **(a-d)** represent the switching rates of the dorsal default mode network (DDMN), ventral default mode network (VDMN), anterior salience network (ASAL), and global switching rate across groups, respectively. **(e-h)** show the association between switching rate of these networks and the mean heart rate (HR) across groups.

**Table 1.**
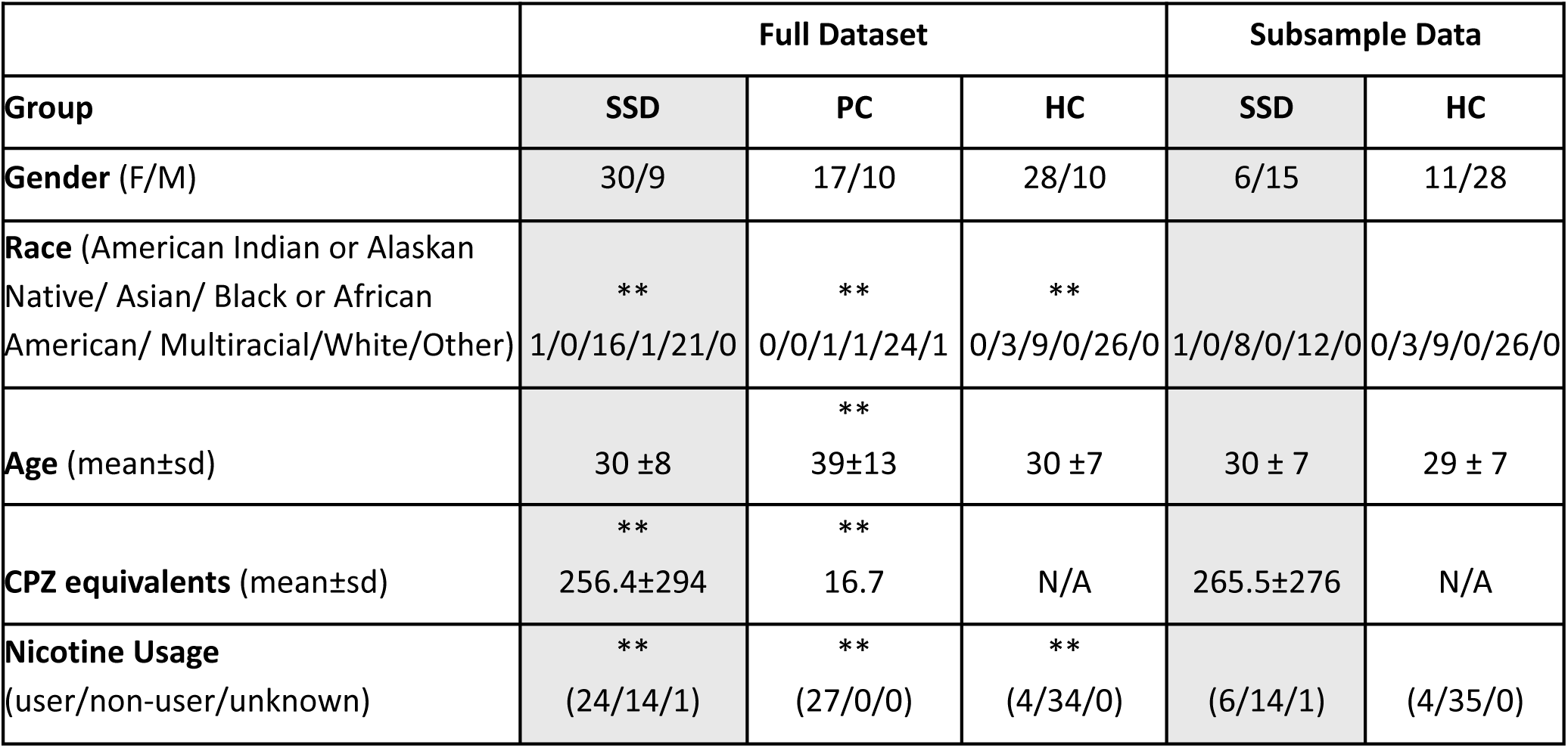
Group demographics. SSD: participants with schizophrenia spectrum disorder, PC: psychiatric controls, HC: healthy controls, F: female, M: male, sd: standard deviation, CPZ: chlorpromazine. (**) denotes p <0.001, when ** is across all groups this indicates a group difference between all comparison of SSD, PC, or HC. The ** for PC in age reflects age was higher for PC compared to both SSD and HC however HC and SSD had no group differences.

**Table 2.** Group Comparisons of Switching Rate Across Networks. Networks include Dorsal Default Mode (DDMN), Ventral Default Mode (VDMN), Anterior Salience Network (ASAL), and Global Switching Rate (Global). Groups include Schizophrenia Spectrum Disorder (SSD), Healthy Controls (HC), and Psychiatric Controls (PC). Reported values represent differences in group means (Diff), lower bounds of the 95% confidence intervals (Lower_CI), and upper bounds of the 95% confidence intervals (Upper_CI).

| Network | Comparison | Diff | Lower_CI | Upper_CI | p-adj |
| --- | --- | --- | --- | --- | --- |
| DDMN | SSD-HC | -1.40E-03 | -5.74E-03 | 2.94E-03 | 0.73 |
| DDMN | PC-HC | 3.36E-03 | -1.08E-03 | 7.80E-03 | 0.18 |
| DDMN | PC-SSD | 4.76E-03 | 6.39E-04 | 8.87E-03 | 0.02 |
| VDMN | SSD-HC | 3.17E-03 | -9.82E-04 | 7.32E-03 | 0.17 |
| VDMN | PC-HC | 4.11E-03 | -1.35E-04 | 8.36E-03 | 0.06 |
| VDMN | PC-SSD | 9.43E-04 | -3.00E-03 | 4.88E-03 | 0.84 |
| ASAL | SSD-HC | -2.25E-04 | -4.73E-03 | 4.28E-03 | 0.99 |
| ASAL | PC-HC | 5.71E-03 | 1.10E-03 | 1.03E-02 | 0.01 |
| ASAL | PC-SSD | 5.93E-03 | 1.66E-03 | 1.02E-02 | 0.004 |
| Global | SSD-HC | 1.53E-03 | 1.51E-04 | 2.92E-03 | 0.03 |
| Global | PC-HC | 3.45E-03 | 2.04E-03 | 4.87E-03 | <0.001 |
| Global | PC-SSD | 1.92E-03 | 6.10E-04 | 3.23E-03 | 0.003 |

### Group Differences in the Relationship Between Switching Rate and Mean Heart Rate or Nicotine

We tested whether mean heart rate interacted with group status (PC, SSD, HC) to predict switching rate across multiple networks (DDMN, VDMN, ASAL, Global) **(Figure 2e-h)**. None of the interactions were significant (all *p* > 0.4), indicating that the relationship between mean HR and switching rate did not differ by group in any network. When we applied the same methods to test if nicotine usage interacted with group to predict switching across these networks, we found a significant interaction between nicotine usage and group on VDMN switching rate **(Supplementary Figure 1)**. This analysis included only HC and SSD groups, as all PC participants were nicotine users. HC non-users had lower switching rates than HC users (β = –0.0128, SE = 0.0043, t(126) = –3.00, p = 0.0032, q=0.03), whereas SSD non-users did not differ from SSD users, resulting in a trend toward nicotine × SSD interaction (though not significant after multiple comparison corrections; β = 0.0130, SE = 0.0049, t(126) = 2.63, p = 0.0097, q=0.07). This pattern suggests that the relationship between nicotine usage and switching rate is modulated by clinical groups, with nicotine use associated with a higher VDMN switching rate in HC participants, but no significant effect in SSD participants.

### Group Comparison of Cognitive Performance

We found SSD participants had significantly lower performance during the 3-digit CPT condition than HC participants (*t*(42)=3.54, *p* < 0.001) (**Figure 3a**).

**Figure 3.**
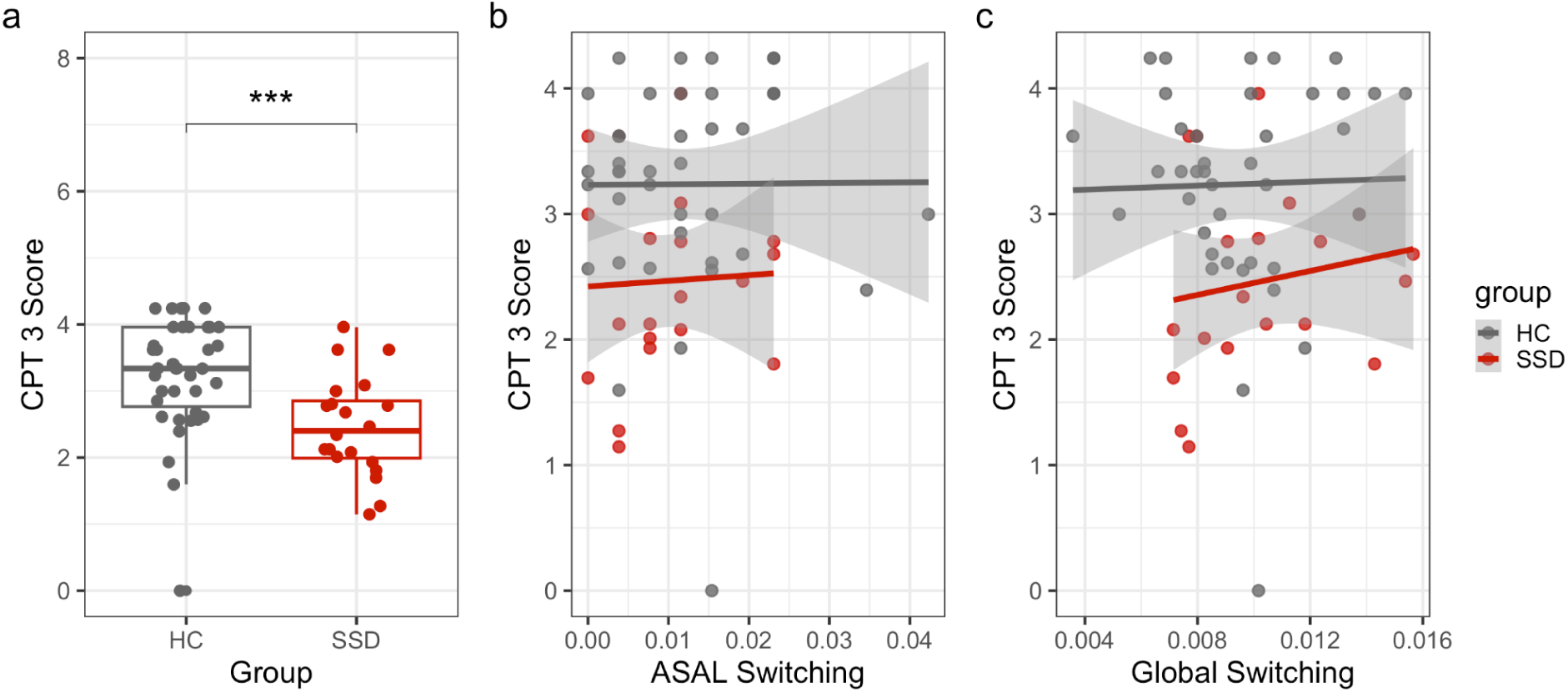
Relationship between cognitive performance and network switching. **a)** Group comparison of continuous performance task scores at the medium level difficulty (CPT 3 Score) for healthy controls (HC) in grey, and participants with schizophrenia spectrum disorder (SSD) in red. The relationship between switching rate and CPT 3 Score shown for **b)** anterior salience network switching (ASAL) and **c)** global switching.

### Relationship between Network Switching and Cognitive Performance

We tested if the global switching rate, and the switching rate of the anterior salience network, related to cognitive task performance in either group; we found no significant relationships (HC-Global: β = –24.14, SE = 73.7, *t* =-0.33, *p* = 0.7; HC-ASAL: β =0.96, SE = 14.70, *t* = 0.07, *p* = 0.95; SSD-Global: β = –24.1, SE = 73.7, *t* =-0.33, *p* = 0.75; SSD-ASAL: β =-0.39.5, SE =28.3, *t* = –1.40, *p* = 0.19). Additionally, there were no group interactions in our test to see whether network switching was related to cognitive performance (Global: β = 2.55, SE = 101.4, *t* =0.03, *p* = 0.98; ASAL: β = –33.1, SE = 37.8, *t* =-0.89, *p* = 0.38).

### Heart Rate Moderates the Relationship Between Network Switching and Performance in Attention in Healthy Controls but Not in Schizophrenia Spectrum Disorder

We next examined whether autonomic arousal moderated the relationship between network switching and cognitive attention. There was no three-way interaction among groups, global switching, and heart rate (β = −1191.3, p = 0.05). Similarly, no three-way interaction among groups, ASAL switching, and HR reached significance. However, when we conducted separate moderation analyses for each group, HR significantly moderated the relationship between global switching and task performance in HC participants (β = 747.1, SE = 33.3, t = 2.24, q = 0.03). Specifically, at higher arousal levels, greater global switching was associated with better task performance. In contrast, at lower arousal levels, greater switching was associated with worse performance. In contrast, no significant moderation was observed in SSD participants (β = –97.5, SE = 585.1, *t* = –0.17, *p* = 0.87). A similar pattern was observed for ASAL switching, with a significant moderation of mean HR in HC (β = 248, SE = 91.2, *t* = 2.72, *p* = 0.01) but not SSD (β = 144.4, SE = 176, *t* = 0.82, *p* = 0.43) participants (**Figure 4**).

**Figure 4.**
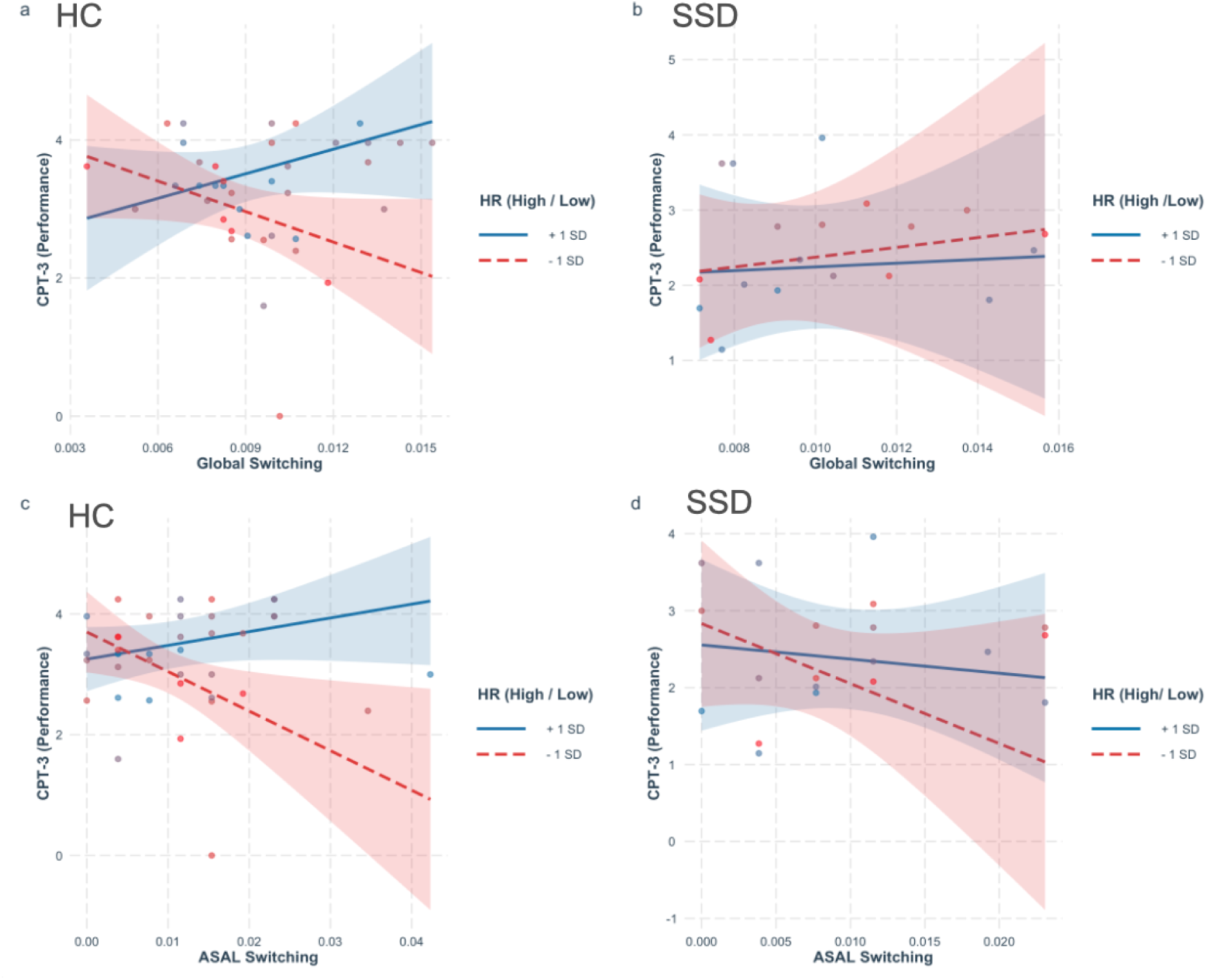
Mean Heart Rate Moderation of Network Switching and Cognitive Performance. Interaction plots showing that mean heart rate (HR) moderates how global switching and anterior salience network (ASAL) switching relate to CPT-3 scores in HC (**a,c)** but not in patients with SSD **(b,d)**. Red represents low heart rate (more than one standard deviation below the mean), and blue represents high heart rate (more than one standard deviation above the mean).

## Discussion

Cognitive deficits are a core feature of schizophrenia, yet their neural underpinnings remain poorly understood. Here, we sought to identify whether a potential neural marker, network switching — a measure associated with cognition (Bassett et al., 2011; Braun et al., 2015; Pedersen et al., 2018) as well as dependent on arousal state (Kundert-Obando et al., 2026) — is disrupted in schizophrenia, and whether such disruptions relate to attentional deficits. In the combined sample, we did not observe significant group differences in autonomic arousal. However, the psychiatric controls (PC) exhibited higher switching of certain subnetworks compared to the schizophrenia spectrum disorder (SSD) group. Additionally, in a subsample with available attentional measures, we observed nominal group differences in global switching rate, as well as evidence that HR moderated the relationship between global switching and attentional task performance in healthy controls (HC), but not SSD, participants. Taken together, these findings provide preliminary evidence that switching rate differentiates PC from SSD and that arousal’s modulatory role in the relationship between global switching and task performance is disrupted in SSD. This disruption may reflect underlying mechanisms that may contribute to cognitive impairments in schizophrenia.

Prior studies have suggested that autonomic arousal is disrupted in schizophrenia (Clamor et al., 2014, 2019; Schlier et al., 2019). Specifically, patients with schizophrenia tend to exhibit a higher mean HR and lower HRV. In the present study, we were unable to identify significant differences in autonomic arousal measures across our groups, possibly due to small sample sizes. However, visual inspection of the results (**Figure 1**) suggests that our results trend towards the direction reported in prior findings. One potential explanation for these small effect sizes may also relate to characteristics of our HC sample. In the general population, the mean HR is typically around 60 beats per minute (bpm); however, in our healthy controls group, several participants (N=66%) exhibited mean HR above 60 bpm. This may have reduced our ability to detect significant group differences in heart rate.

Additionally, we investigated whether subnetworks of the default mode network (DMN) and the anterior salience network (ASAL) exhibited altered switching rates across groups, given prior evidence that these networks show arousal-dependent switching rates (Kundert-Obando et al., 2026). This investigation allowed us to determine whether arousal-dependent neural markers are disrupted in PC or SSD. We hypothesized that, in participants with SSD, the switching rate of the ASAL network would be higher and DMN switching lower, consistent with prior reports (Braun et al., 2015; Gifford et al., 2020; Li et al., 2025). However, we found no group differences between SSD and HC. This result is not uncommon, as other studies have similarly reported no group differences in salience and default mode network switching in SSD participants compared to HC participants (Wei et al., 2022).

Interestingly, we found that the PC group exhibited higher ASAL switching compared to both the HC and SSD groups, as well as higher DDMN switching compared to the SSD group. These findings support our hypothesis that switching rate would differentiate PC from SSD, as previous studies have shown that different psychiatric conditions are associated with distinct switching rate patterns (Han et al., 2020). While one might hypothesize that these differences are attributable to arousal differences between PC and SSD, we did not observe significant group differences in autonomic arousal measures; therefore, we cannot make this claim.

One potential explanation is that the PC group was composed exclusively of nicotine users. In the HC and SSD groups, we tested whether nicotine use moderated the relationship between clinical diagnosis and switching rate; however, nicotine use influenced switching rate only in the HC group. Because we did not include non-nicotine users in the PC group, future research should examine whether switching rates differ among individuals with psychiatric symptoms who use nicotine compared to those who do not. Another possible explanation relates to evidence that switching rate is associated with the gene expression of several neurotransmitter systems (Zhen et al., 2025). Given that neurotransmitter expression may relate to PC and SSD in complex ways, future research could investigate whether pharmacological agents targeting specific neurotransmitter systems or brain stimulation (Webler et al., 2026) produce differential network switching patterns in PC and SSD.

Further, we investigated whether the switching rates of DMN subnetworks and ASAL would relate to mean HR and if such relationships were altered across our groups. We found no significant relationship between network switching and autonomic arousal, nor did this relationship differ between groups. This null finding may reflect a limitation of mean HR as an index of arousal, particularly given that HR did not significantly differ between groups. Future studies should examine whether alternative measures, such as pupillometry or electroencephalographic recordings, more effectively capture continuous arousal fluctuations that relate to network switching dynamics. Additionally, future studies should examine whether specific brain regions, rather than large-scale networks, more directly reflect disruptions in arousal-related switching in SSD. For example, our recent work in healthy controls indicated that the thalamus exhibits robust arousal-dependent switching rates (Kundert-Obando et al., 2026).

Next, we hypothesized that arousal would moderate the relationship between global brain switching rate and task performance in HC, and that this interaction would be absent in SSD. Additionally, given evidence that autonomic arousal is closely linked to the salience network (Wei & Wu, 2021), we conducted an exploratory analysis to test whether autonomic arousal would moderate the association between ASAL dynamics and task performance in HC, but not in individuals with SSD. Both hypotheses were supported when conducting the moderation analyses by group (HC versus SSD) for both global and anterior salience network switching. These findings replicate prior work demonstrating that arousal moderates the relationship between global switching and cognitive task performance in HC in attention (Kundert-Obando et al., 2026), and they provide preliminary evidence that this interaction is disrupted in SSD.

A possible explanation for the absence of this interaction in SSD may be attributed to the effects of antipsychotic medication. Braun et al. (2016) reported that healthy controls who were administered 120 mg of dextromethorphan, used as an NMDA receptor antagonist to test for glutamate’s role in network disruption, exhibited switching rate patterns similar to those observed in individuals with schizophrenia. One caveat is that this study reported elevated switching rates across all networks in participants with and without schizophrenia who received dextromethorphan, a pattern that has not been consistently replicated. Nonetheless, future studies should determine whether unmedicated patients also exhibit disrupted arousal–network associations with an array of cognitive task performance measures. Overall, these findings imply that global switching, as well as anterior salience network switching, may represent neural targets for regulating brain–arousal interactions that may support cognitive performance of attention in schizophrenia.

In conclusion, our findings provide evidence that switching rate patterns differ in SSD compared to our psychiatric controls. Additionally, we replicate prior work suggesting that global switching is an arousal-dependent neural marker related to cognitive task performance in attention, and extend these findings to the anterior salience network. Further, we provide initial evidence that this arousal–network interaction is absent in schizophrenia spectrum disorder and may contribute to specific aspects of cognitive impairment, like attention, that are often reported in schizophrenia. Replication in larger samples is needed to determine whether arousal–switching dynamics represent viable clinical targets for improving cognitive performance in schizophrenia.

## Data Availability

All data produced in the present study are available upon reasonable request to the authors.

## Data and Code Availability Statement

This study used data acquired from Vanderbilt University Medical Center. If interested in acquiring this data, please reach out to Dr. Heather Ward at and Dr. Julia Shieffield at. Code for reproducing this study will be available on https://github.com/krogge-obando/net_switch_psychosis.

## Acknowledgments

The authors would like to acknowledge the team of the Vanderbilt Psychotic Disorders Program that was instrumental in the recruitment of participants in this study. Parts of this work were presented in abstract form at the 2026 Society of Biological Psychiatry meeting.

## Authors’ contributions

KKO–conceptualization, formal analysis, investigation, project administration, visualization, Writing-original draft, Writing – review & editing; ARK--data curation, investigation, methodology, project administration, writing-review-editing; SW – conceptualization, investigation, methodology; HP-methodology, writing-review-editing; EP-methodology; AM-validation; BPR-data curation, methodology; SP-Writing-review-editing; JMS–resources, investigation, writing-review-editing, project administration; HBW--resources, investigation, writing-review-editing, project administration; CC-resources, investigation, writing-review-editing, project administration

## Supplementary Information

**Supplementary Figure 1.**
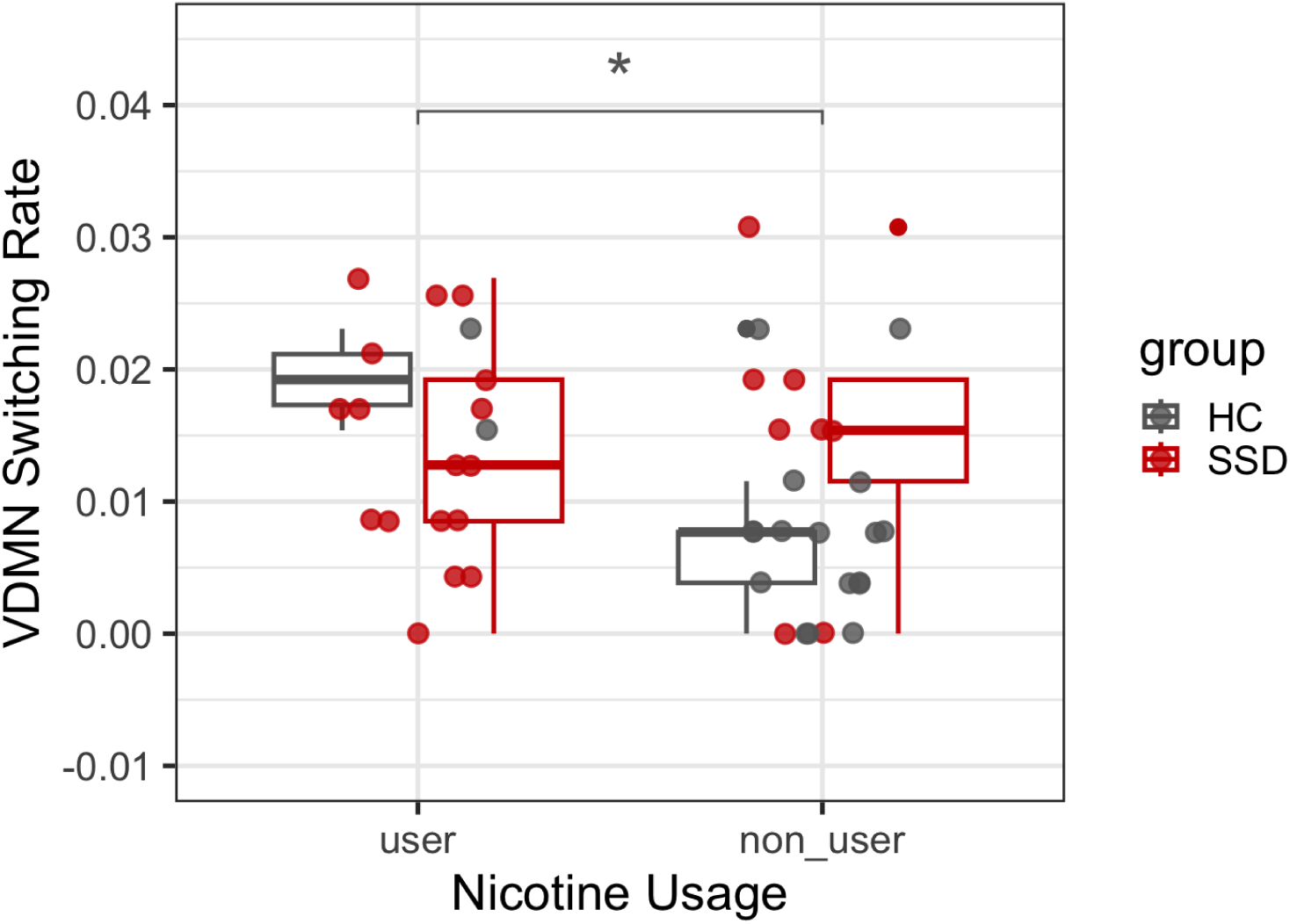
VDMN Switching Across Nicotine Users and Non-Users Across Groups. A significant difference was found only in the HC group. (*) p<0.05.

